# High versus low dose of 14 days treatment of primaquine in *Plasmodium vivax* infected patients in Cambodia: a randomised open-label efficacy study

**DOI:** 10.1101/2025.01.01.25319862

**Authors:** Virak Eng, Dysoley Lek, Sitha Sin, Lionel Brice Feufack-Donfack, Agnes Orban, Jeremy Salvador, Dynang Seng, Sokleap Heng, Nimol Khim, Kieran Tebben, Claude Flamand, Cecile Sommen, Rob W. van der Pluijm, Michael White, Benoit Witkowski, David Serre, Jean Popovici

## Abstract

**Background:** The WHO malaria treatment guidelines recommend a total dose in the range of 3·5 to 7·0 mg/kg of primaquine to eliminate *Plasmodium vivax* (*P. vivax*) hypnozoites and prevent relapses. There are however indications that for tropical *P. vivax* isolates, notably from Southeast Asia, the lower dose of 3·5 mg/kg is insufficient. Determining the most effective regimen to eliminate *P. vivax* hypnozoites is needed to achieve elimination of this malaria parasite.

**Methods:** We conducted an open-label randomised controlled trial in Kampong Speu province, Western Cambodia. *P. vivax* infected patients with uncomplicated malaria, diagnosed at the community level or in health centres of the province, were offered to participate. Patients aged less than 15 years old, and pregnant or breastfeeding women were excluded. Enrolled patients were treated with a blood schizonticidal artesunate regimen of 2 mg/kg/day for 7 days. Upon enrolment, patients’ glucose-6-phosphate dehydrogenase (G6PD) activity was determined. G6PD normal patients were randomly assigned (2:2:1) to receive either (i) 3·5 mg/kg (low dose as 0·25 mg/kg/day) or (ii) 7·0 mg/kg (high dose as 0·5 mg/kg/day) of primaquine over 14 days or (iii) no primaquine as comparator arm. G6PD deficient patients were assigned to the no-primaquine comparator arm. Randomisation was done by blocks of 5 using sealed envelopes. Upon enrolment, patients were relocated to the study site in Aoral town where no malaria transmission occurs to ensure that they were not reinfected during their 90-day follow-up. After 90 days of relocation, G6PD normal patients in the no-primaquine arm were provided 3·5 mg/kg for 14 days of primaquine to be taken unsupervised. At day 90, all the patients returned home and they were further followed monthly for three months until day 180. The primary outcome was the treatment failure rate defined as the proportion of patients with at least one *P. vivax* recurrence within 90 days of relocated follow-up. All patients that completed treatment and complied with relocation without interruption before any recurrence was detected were included in the primary efficacy analysis. All patients enrolled and assigned to an intervention arm were included in the safety analysis. The study is registered on ClinicalTrials.gov (NCT04706130).

**Findings:** Between Nov 10, 2021, and Feb 10, 2024, a total of 160 patients were enrolled and 156 were allocated to one of the three study arms. Of these, 37 G6PD deficient patients were assigned to the no primaquine arm and 119 G6PD normal patients were randomised: 24 in the no primaquine arm, 49 in the primaquine 3·5 mg/kg arm, and 46 in the primaquine 7·0 mg/kg arm. The proportion of participants with at least one *P. vivax* recurrence within 90 days in the no primaquine arm was 81·4% (95% CI 69·6-89·2). The proportion of participants with recurrence was higher in the low dose primaquine arm (24·4%, 95% CI 14·2-38·7) compared to the high primaquine arm (4·7%, 95% CI 0·8-15·5, p=0·0141) resulting in a hazard ratio of high dose primaquine compared to low dose of 0·17 (95% CI 0·04-0·79, p=0·0229). Both primaquine arms were well tolerated.

**Interpretation:** Not providing primaquine to patients led to a considerable rate of *P. vivax* recurrence. The risk of *P. vivax* recurrence was 5·9 times lower for the 7·0 mg/kg of primaquine treatment compared to 3·5 mg/kg. Tolerability and safety of both primaquine regimens in G6PD normal individuals was comparable. Policy makers in Cambodia and most likely in other Southeast Asian countries should endorse the 7·0 mg/kg of primaquine regimen to reduce the risk of *P. vivax* relapses.

**Funding:** National Institutes of Health (R01AI146590)

**Research in context:** *Evidence before this study:* The WHO treatment guidelines for preventing *Plasmodium vivax* relapses using primaquine recommend a range of 3·5-7·0 mg/kg. These guidelines mention that for *P. vivax* infections acquired in Southeast Asia and Oceania, 7·0 mg/kg should be preferred. We searched Pubmed for randomized controlled trials studies containing the terms “vivax” and “primaquine” published between 1990 and November 2024, with no language restrictions, to identify studies comparing primaquine regimen to treat *P. vivax* infections. Our search retrieved only two studies comparing head-to-head the efficacy of 3·5mg/kg with 7·0 mg/kg primaquine administered over a same duration of 14 days but were both conducted in South Asia and both showed similar efficacy of high dose compared to low dose primaquine. No studies compared 3·5 and 7·0 mg/kg of primaquine administered over 14 days in South East Asian countries.

*Added value of this study:* This randomised controlled trial compared the efficacy of a total dose of 3·5 mg/kg and 7·0 mg/kg of primaquine over 14 days to prevent relapses of *P. vivax* in Cambodia. Our study design minimized confounding factors affecting therapeutic efficacy evaluation. This study confirms that 7·0 mg/kg has greater efficacy to prevent recurrences than the low 3·5mg/kg regimen, with comparable safety and tolerability in G6PD normal patients from Cambodia.

*Implications of all the available evidence:* Our results confirm that 7·0 mg/kg of primaquine should be recommended to prevent relapses in *P. vivax* infections acquired in Cambodia and most likely in South East Asia. National treatment guidelines in those countries should be changed to endorse this regimen rather than the 3·5 mg/kg.

## Introduction

*Plasmodium vivax* is the parasite responsible for the majority of malaria cases outside Africa, with more than three billion people living within *P. vivax* transmission limits.^1^ Upon mosquito inoculation, a fraction of the injected parasites remain dormant in human hepatocytes as hypnozoites and can reactivate after the initial infection causing relapses.^2,3^ Repeated relapses of *P. vivax* cause significant morbidity and mortality in individuals living in endemic areas.^4–6^ In addition, the hypnozoite reservoir of *P. vivax* make this parasite resilient to elimination efforts and the proportion of *P. vivax* malaria tends to increase during malaria elimination campaigns.^7–12^ In many endemic countries, *P. vivax* is now the most prevalent malaria parasite and dedicated strategies are needed for its elimination.^13^

Only 8-aminoquinolines such as primaquine and more recently tafenoquine, are able to eliminate hypnozoites. Despite primaquine being used for decades, the most appropriate regimen for achieving complete elimination of hypnozoites is still unclear (referred to as radical cure when combined to a schizonticide drug killing blood-stage parasites). The WHO malaria treatment guidelines recommend to administer 0·25 to 0·5 mg/kg/day of primaquine over 14 days (total dose of 3·5 to 7·0 mg/kg) to achieve radical cure, and state that tropical *P. vivax* prevalent in Southeast Asia and Oceania should be treated with a total dose of 7·0 mg/kg rather than 3·5 mg/kg.^14^ Nonetheless, the guidelines also acknowledges that “No direct comparison has been made of higher doses (0·5 mg/kg body weight for 14 days) with the standard regimen (0·25 mg/kg body weight for 14 days)”. A number of trials conducted in South East Asia and Oceania evaluated different primaquine doses administered over variable durations^15–18^, but none compared 3·5 and 7·0 mg/kg total dose of primaquine administered over 14 days. Two studies compared 3·5 and 7·0 mg/kg of primaquine over 14 days, but both were conducted in India and not in South East Asia or Oceania^19,20^. Both concluded similar efficacy between the two primaquine regimens. To improve compliance of primaquine treatment, the WHO recently added a recommendation to administer a total of 3·5 mg/kg over 7 days instead of 14 days.

Many endemic countries have been reluctant to roll out primaquine, especially at the high 7·0 mg/kg total dose, due to safety concerns in glucose-6-phosphate dehydrogenase (G6PD) deficient individuals in whom primaquine can cause severe haemolytic anaemia.^21,22^ For many endemic countries where G6PD deficiency is common, such as in Cambodia and in general in Southeast Asia, the treatment guidelines require that the G6PD status of an individual is determined prior to administration of primaquine and most countries recommend a total primaquine dose of 3·5 mg/kg.^22,23^ It is important to rigorously evaluate the efficacy of the different primaquine regimens for radical cure treatment as substandard treatment will result in relapses within the same individual but will also contribute to ongoing transmission in the population, thereby hindering malaria elimination efforts.

*P. vivax* drug efficacy studies are complicated since, following initial treatment, parasite recurrences can arise from blood-stage parasite recrudescence, reinfection from a new mosquito inoculation or relapses from hypnozoite reactivation.^24,25^ In this study, we overcome this difficulty by relocating patients from a malaria endemic region to a non-endemic area. We report the results of a randomised controlled trial evaluating the efficacy of 3·5 versus 7·0 mg/kg total dose over 14 days of primaquine to prevent *P. vivax* recurrences.

## Methods

### Study design

We conducted an open-label randomised controlled trial to compare the efficacy of low (3·5 mg/kg total dose as 0·25 mg/kg/day for 14 days) and high dose (7·0 mg/kg total as 0·5 mg/kg/day for 14 days) primaquine in G6PD normal *P. vivax* infected individuals. The study was conducted in our field site located in Aoral town, Kampong Speu Province, Western Cambodia. The protocol was approved by the National Ethics Committee for Health Research of Cambodia (158-NECHR, 06/29/2020) and by the University of Maryland IRB (HP-00091095). The study was overseen by the NIH DMID (Protocol 20-0010)

### Participants

Patients aged more than 15 years old, able to provide consent and willing to participate, seeking treatment and diagnosed with an acute symptomatic *P. vivax* infection by rapid diagnostic test at the community or health centre level were eligible for inclusion. Exclusion criteria were: aged less than 15 years old, sign of severe malaria, haemoglobin concentration below 8·0 g/dL, pregnant and breastfeeding women, known sensitivity to the study drugs and intake of antimalarial drug in the past month prior to inclusion. Since prolonged artesunate treatment, as administered in this study prior to primaquine, has been shown to possibly lead to neutropenia^26^, initially patients were excluded if they presented with a neutrophil count of less than 1·5×10^9^ cells/L. However, after approval from the Safety Monitoring Committee of the study an amendment to the study protocol was made to remove this exclusion criterion and instead monitor clinical signs of neutropenia. Only PCR-confirmed *P. vivax* mono-infections were initially eligible. However, we amended the study protocol to include mixed *P. vivax* with other *Plasmodium* species infections detected by PCR to ensure faster enrolment rates. Only patients with a G6PD activity of more than 4 U/gHb for males or 6 U/gHb for females were eligible for primaquine administration according to the Cambodian National Treatment Guidelines. G6PD activity was measured by spectrophotometry (Trinity Biotech quantitative G6PD assay, adapted for use on the Integra 400 analyzer, Roche Diagnostics). Patients with G6PD levels below these thresholds were assigned to the no-primaquine arm. All patients or their guardians provided written informed consent and written assent was obtained for all patients aged 15-18 years old.

### Randomisation and masking

Enrolled patients were treated with 2 mg/kg/day of artesunate for seven days (total dose 14 mg/kg). Males with a G6PD activity below 4 U/gHb and females with activity below 6 U/gHb were allocated to the no primaquine comparator arm. Male and female patients with G6PD values above these thresholds were randomly assigned to either low dose primaquine (0·25 mg/kg for 14 days, total dose 3·5 mg/kg), high dose primaquine (0·5 mg/kg for 14 days, total dose 7·0 mg/kg) or no primaquine, in a 2:2:1 ratio. Randomisation was done using sealed envelopes in blocks of five and was performed by an independent team member who communicated the allocation to the clinician on-site after screening and enrolment was completed. Randomization was performed in all cases within the first 6 days of artesunate treatment so all participants randomized to the primaquine arms started their treatment on day 7. The clinician on-site was responsible for the enrolment of patients in the study. All treatments were open labelled.

### Procedures

All patients started the blood schizonticidal artesunate regimen (RT Nate, Bonn Schtering Bio Sciences, India) on the day of inclusion before the screening results were available. This artesunate regimen was adapted from a previous study to ensure complete elimination of blood-stage parasites while being rapidly eliminated and not providing post-treatment prophylaxis that could mask failure of primaquine.^27^ In case the screening led to an exclusion of patients, artesunate regimen was interrupted and a complete artesunate-mefloquine 3-day regimen was provided following the Cambodian National Treatment Guidelines. Excluded patients were then referred to the Aoral Health Centre for G6PD activity determination and primaquine administration. For enrolled patients, all treatments were directly supervised at all times. Artesunate treatment was provided at 2 mg/kg/day for seven days from day 0 to day 6 (appendix Table S1). Following results of screening and arm allocation, patients in the two primaquine arms started primaquine (Remedica, Ltd., Cyprus) on day 7 after the last dose of artesunate. Patients receiving primaquine were treated daily for 14 days from day 7 to day 20 with either low dose (0·25 mg/kg/day) or high dose of primaquine (0·5 mg/kg/day, appendix Table S2). Only antipyretic treatment (paracetamol) was provided concomitantly to some patients at the discretion of the clinical physician.

Upon enrolment, a physical examination was performed. A venous blood sample was collected before treatment to perform a full blood count and determine G6PD activity. From the day of inclusion, patients remained relocated for 90 days in our study site located in Aoral, a malaria transmission-free town. Patients could go in and out of the study site during day time but had to spend the night there to ensure they do not travel to remote forested areas during the follow-up. Clearance of blood-stage parasites was monitored by capillary blood collection 1h, 2h, 4h, 8h, 16h, 24h after the first dose and then daily from day 2 to day 6. Follow-up for patients not receiving primaquine was then done every 48h until day 90 and the end of relocation. Patients receiving primaquine had daily follow-up during the 14 days of primaquine treatment after which follow-up was performed every 48h until day 90. At each follow-up, a physical examination was performed, any adverse event (AE) was recorded and a capillary blood sample was collected by fingerprick into an EDTA microtainer tube. From each sample, thick and thin smears were prepared on microscopy slides for Giemsa staining (3%, 45 min) and 50μL of blood was used for DNA extraction and qPCR detection of malaria parasites performed in the following 24h.

During the follow-up, in case of *P. vivax* recurrence detected by qPCR and confirmed by microscopy, patients were re-treated with the same artesunate regimen received upon inclusion (14 mg/kg over 7 days) but not primaquine. Follow-up was then resumed until the end of the 90 days of relocation. On the last day of relocation, G6PD normal patients who did not receive primaquine were provided the standard 14-day primaquine treatment of 0·25 mg/kg/day, to be taken unsupervised. Participants were further followed for 3 additional monthly visits on day 120, day 150 and day 180 for capillary blood collection.

### Outcomes

The primary outcome of the study was the proportion of patients experiencing at least one *P. vivax* recurrence during the relocated 90-day follow-up. Secondary outcomes included the proportion of patients with *P. vivax* recurrence detected after the end of relocation on day 180, the total number of recurrences occurring during (i) the 90-day relocation (ii) the 180-day complete follow-up, the time to the first *P. vivax* recurrence and clearance of blood-stage parasites following artesunate treatment.

Safety assessment and grading of adverse events (AE) were performed following the NIAID/NIH DMID toxicity table (Appendix Table S3). A clinical examination was performed every 24 or 48h during the 90-day follow-up. Haematological AEs and other drug related AEs that are the known expected risks associated to artesunate and primaquine treatments were particularly monitored. Neutrophil counts were determined before and 7 and 14 days after any artesunate treatment. Haemoglobin concentration (using a HemoCue) and methaemoglobin percentage (measured using a Masimo oximeter) were recorded daily over the course of any treatment and then every 48h during the 90-day follow-up.

### Sample size

We anticipated that at least 50% of patients would have *P. vivax* recurrence during the 90 days follow-up in absence of any radical cure^27,28^. Assuming an efficacy of 92% for the 0·5 mg/kg/day and 73% for the 0·25 mg/kg/day, a Type I error of 5% and a power (1-β) of 90%, a sample size of 80 individuals per arm was required (total 160) to detect statistically significant differences in therapeutic efficacies (target effect) ^27,29,30^. With a 20% loss during follow-up estimated, 100 patients per primaquine arm (total 200) was defined. For the no primaquine arm, assuming a Type I error of 5%, a power (1-β) of 90% and a loss to follow-up rate of 20%, a sample size of 60 patients in this arm (expected ∼ 30 patients with relapses and ∼30 patients without) was defined to provide sufficient power. This arm was not intended as therapy for practice, but aimed to serve as comparator as well as to study relapses in vivo.

### Statistical analyses

Difference in efficacy of the two primaquine regimens was assessed by two-sided Fisher’s exact test using the per protocol population that excluded participants who did not complete the treatment (either because of mixed infections or withdrawal of consent) or interrupted relocated follow-up before any recurrence was detected. The intention-to-treat population was used in a sensitivity analysis where all dropouts were imputed either as treatment success or treatment failure. These analyses were complemented by comparing Kaplan-Meier recurrence-free survival using log-rank analysis. All participants assigned to one of the three study arms were included in the Kaplan Meier analysis. Patients were censored in the Kaplan-Meier analysis at the day of the following events: withdrawal of consent (with or without discontinuation of study drug), mixed *Plasmodium* infections (until this exclusion criterion was removed) and loss to follow-up. Before it was removed from exclusion criteria, in case of mixed infection during the follow-up, patients were referred to the local health centre to be treated with first-line treatment for malaria (artesunate-mefloquine). Interruption of relocation (absence at night from the study site) was tolerated if it lasted less than 4 days (two consecutive 48h follow-up time points). Patients were advised that breach of relocation was not tolerated for more than 4 days and this was considered as withdrawal of consent. No patient had more than 1 relocation interruption during the 90 days follow-up. Missing visits after the 90-day of relocation on day 120, 150 or 180 were all tolerated. Hazard ratios for time to first recurrence were determined by Cox regression. A Prentice-Williams-Peterson (PWP)^31,32^ extended Cox regression analysis was used to compare risks of all recurrences between the three arms taking into account repeated P. vivax recurrences. PWP model stratifies the analysis by the order of events, allowing the baseline hazard to vary for each event. Differences in the number of recurrences, the time to first recurrence and the time to any recurrence (i) between randomized and non-randomized patients in the no primaquine arm and (ii) between low dose primaquine and high dose primaquine arms, were all assessed by Mann Whitney U test. All tests were two-sided. All patients enrolled and assigned to a study arm were included in the safety analysis. All analyses were done using R V12.1 or GraphPad Prism V10.3. There was a Safety Monitoring Committee for this study with pre-planned annual meetings. The study is registered on ClinicalTrials.gov (NCT04706130).

## Results

Between Nov 10, 2021, and Feb 10, 2024, 312 patients were screened for inclusion, of whom 129 declined to participate, 23 did not meet inclusion criteria and 160 were enrolled in the study, of whom four were later excluded as they were not filling laboratory screening criteria (figure 1). All patients were Cambodian. Enrolment was halted following consultation with the Safety Monitoring Committee despite not have reached the initially planned sample size as the study reached its end date and concomitantly with decrease in malaria transmission and enrolment rates in the study area. Of the 156 enrolled patients, 37 (23·7%) had a G6PD activity below the thresholds for primaquine administration and were assigned to the no-primaquine arm. The 119 remaining G6PD normal patients were randomly assigned to one of the three arms: 49 in the low-dose primaquine arm, 46 in the high dose and 24 in the no primaquine arm. The final sample size was 49, 46 and 61 patients enrolled in the low dose primaquine, high dose primaquine and no primaquine arms respectively. Baseline patients’ characteristics were similar between the three study arms and between G6PD normal patients randomised to the no primaquine arm and G6PD deficient patients assigned to the no primaquine arm (table 1). The majority of enrolled participants were males reflecting the current malaria epidemiology in Cambodia where adult males are the most at-risk individuals due to occupational exposure.^33^ The median parasitemia was 5573 parasites per μL (IQR 3223-10795) in the low dose primaquine arm, 6470 parasites per μL (IQR 2815-12557) in the high dose primaquine arm, 4890 parasites per μL (IQR 2484-16465) in participants randomised to the no primaquine arm and 5976 parasites per μL (IQR 3897-11561) in G6PD deficient participants assigned to the no primaquine arm.

**Figure 1.**
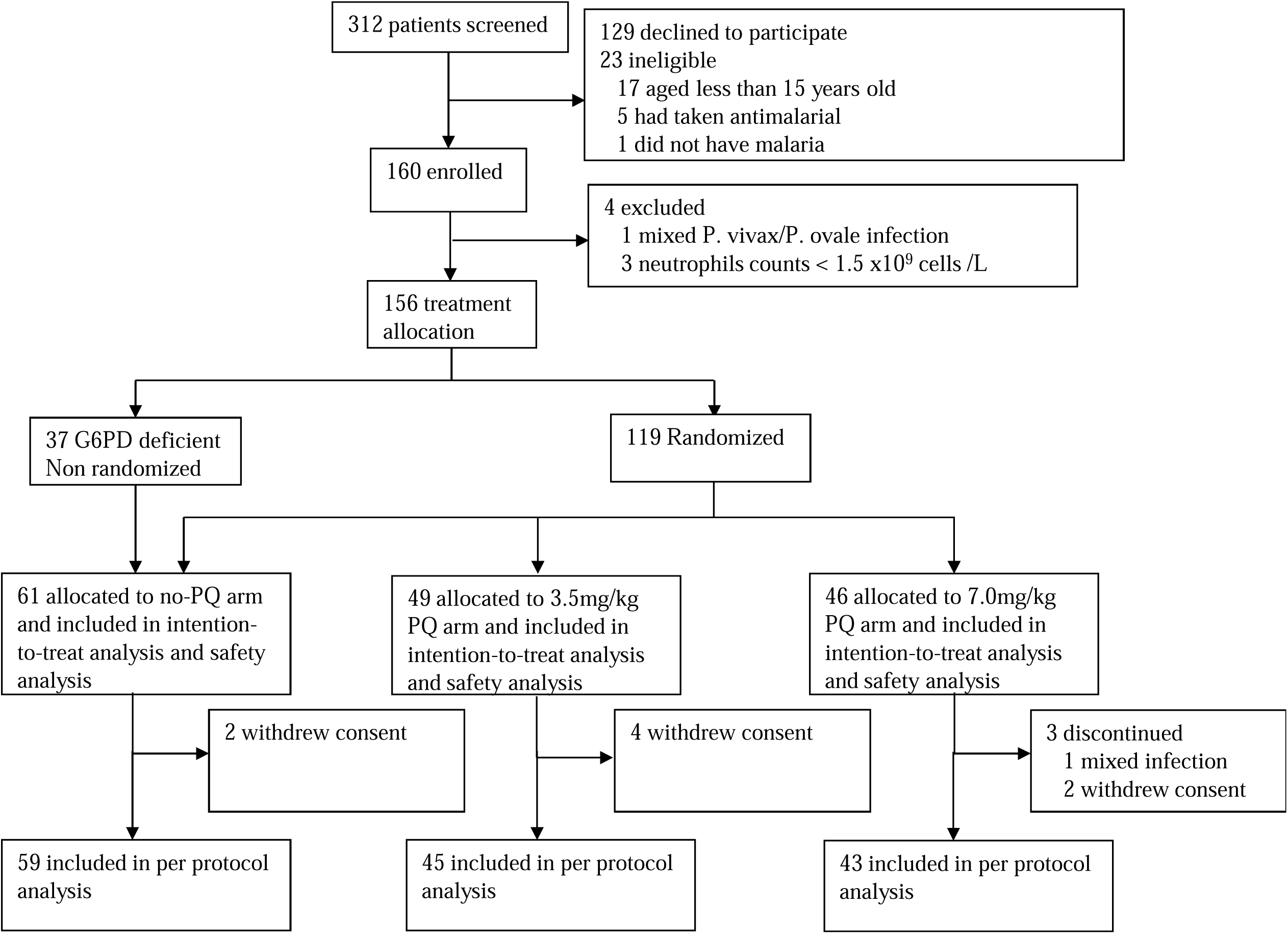
CONSORT diagram. G6PD= glucose-6-dehydrogenase, PQ= primaquine.

In the per protocol population, the proportion of participants with at least one *P. vivax* recurrences detected by day 90 was 81·4% (48/59, 95% CI 69·6-89·2) in the no primaquine arm. There was no statistically significant difference in recurrence rates between randomized G6PD normal patients (77·3%, 17/22, 95% CI 56·6-89·9) and non-randomized G6PD deficient patients (83·8%, 31/37, 95% CI 68·9-92·3, p=0·7309) (table 2). Among those receiving primaquine, 24·4% (11/45, 95% CI 14·2-38·7) of the participants in the low dose primaquine arm had a recurrence, statistically significantly higher than those in the high dose primaquine arm 4·7% (2/43, 95% CI 0·8-15·5, p=0·0141). Efficacy of primaquine obtained here are similar to those that were anticipated. The ITT analysis led to similar results with statistically significantly higher recurrences in the low dose primaquine arm compared to the high dose arm, whether dropouts were imputed as treatment success (p=0·0150) or treatment failure (p=0·0236) (Appendix Table S4). Missing data are listed in Appendix Table S5. Log-rank analysis confirms that *P. vivax* recurrence-free survival by day 90 was statistically significantly different between low and high dose primaquine arms (p=0·0100) (figure 2A). The same results were observed when restricting this analysis to randomized patients only (Appendix figure S1). The hazard ratios (HR) for recurrence compared to no primaquine were 0·14 (95% CI 0·07-0·27) for the low-dose primaquine arm and 0·03 (95% CI 0·01-0·10) for the high-dose arm (p<0·0001). The HR of high dose primaquine compared to low dose was 0·17 (95% CI 0·04-0·79, p=0·0229).

**Figure 2.**
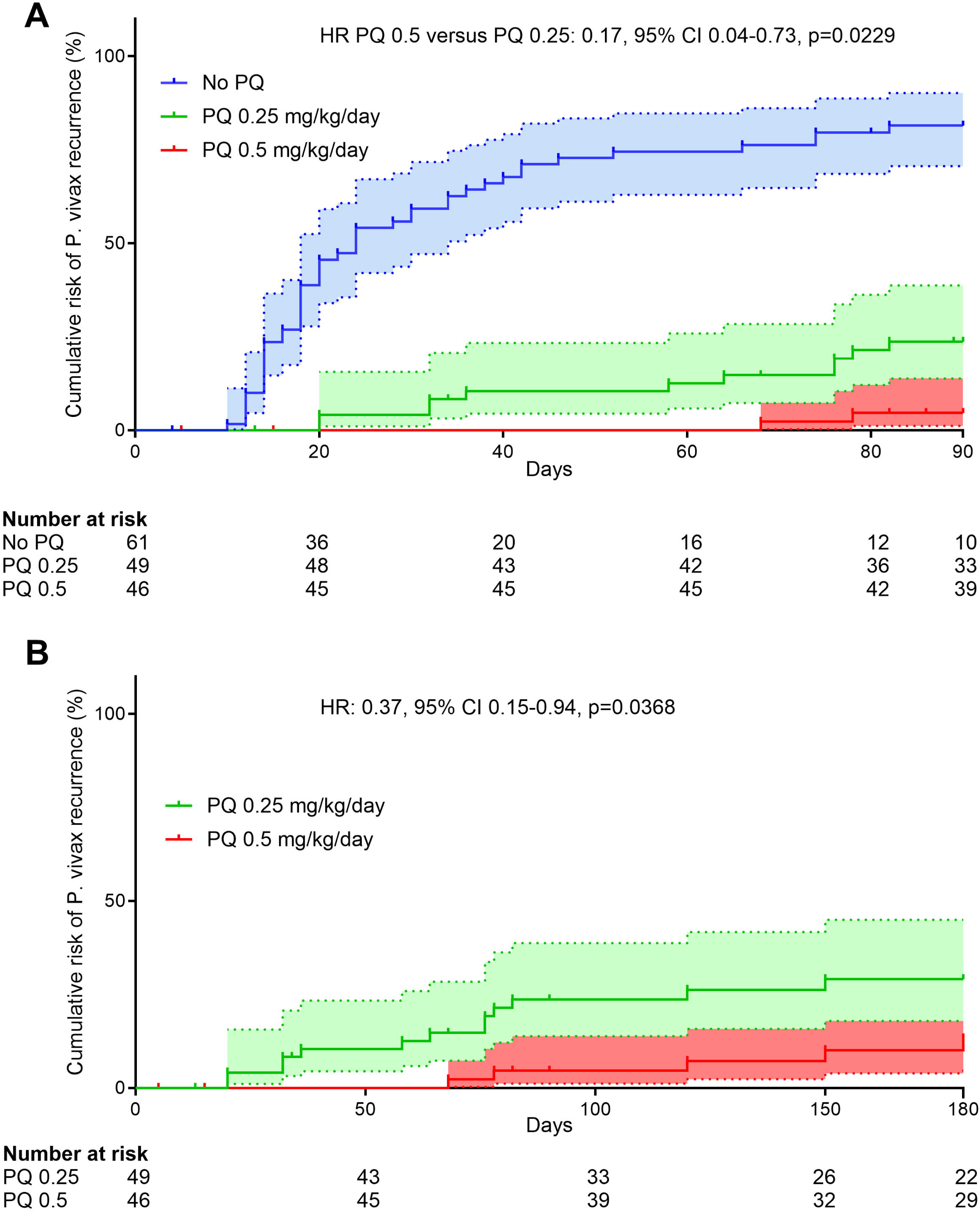
Risk of *Plasmodium vivax* recurrence. Kaplan-Meier graphs showing the risk of *P. vivax* recurrences at day 90 (A) and day 180 (B) for patients treated with artesunate and receiving in addition either no primaquine (No PQ), 14 days of 0·25 mg/kg/day of primaquine (PQ 0·25) or 14 days of 0·5 mg/kg/day of primaquine (PQ 0·5). Shading represents 95% CI.

The Cox model accounting for repeated *P. vivax* recurrence events showed that compared to the no primaquine arm, the risk of recurrence was 4·9 times lower (95%CI 2·49-9·55, p<0·0001) in the low dose primaquine arm, and 36 times lower (95%CI 9·28-142, p<0·0001) in the high dose primaquine arm. The risk of recurrence was 7·5 times lower in the high primaquine arm compared to the low primaquine arm (95%CI 1·63-34·2, p=0·0067).

Between day 90 and day 180, two additional participants in the low dose primaquine arm and three in the high dose primaquine arm experienced their first *P. vivax* recurrences. Recurrence-free survival analysis resulted in incidence risk of 13·2% (95%CI 5·7-29·1) in the high dose primaquine arm, statistically significantly lower than in the low dose primaquine arm (incidence risk, 29·1%, 95%CI 17·9-45·0, HR 0·37 95%CI 0·15-0·94 p=0·0368) (figure 2B). For the no primaquine arm, we analysed separately randomized G6PD normal individuals that were provided primaquine to be taken unsupervised at day 90 and non-randomized G6PD deficient patients who were not provided primaquine at day 90. Recurrence-free survival analysis showed no difference between G6PD normal and deficient individuals with incidence risks of 81·7% (95%CI 63·6-94·3) and 84·2% (70·7-93·8) for G6PD normal and deficient participants, respectively (p=0·8916).

The mean number of recurrences per patient during the 90-day follow-up in the no-primaquine arm was 1·7 (SD 1·2, range 0-4) and was not statistically significantly different between randomized G6PD normal patients (1·8, SD 1·2, range 0-4) and non-randomized G6PD deficient patients (1·8, SD 1·2, range 0-4, p=0·9728). For patients treated with primaquine, statistically significantly more recurrences per patient occurred in the low primaquine arm (0·3, SD 0·7, range 0-3) compared to the high primaquine arm (0·04, SD 0·21, range 0-1, p=0·0193). Similar trends were observed when considering all recurrences occurring until day 180 with a mean of 2·1 (SD 1·4, range 0-4) recurrences in the no primaquine arm and no statistically significant difference between randomized (2·0, SD 1·3, range 0-4) and non-randomized patients (2·2, SD 1·4, range 0-4, p=0·5868). For primaquine-treated patients, there were statistically significantly more recurrences per patient in the low primaquine arm (0·47, SD 0·94, range 0-4) compared to the high primaquine arm (0·11, SD 0·31, range 0-1, p=0·0367).

Among participants with recurrences, the median time from enrolment to the first recurrence was 20·0 days (IQR 14·0-36·5, range 10-150), 64·0 days (IQR 32·0-80·0, range 20-150) and 120·0 days (IQR 73·0-165·0, range 68-180) in the no-primaquine, low dose primaquine and high dose primaquine arms, respectively. In the no primaquine arm, the time to first recurrence was not statistically significantly different between randomized (18 days, IQR 14-30, range 12-120) and non-randomized patients (24 days, IQR 14-38, range 10-150, p=0.4521). The time to first recurrence was statistically significantly shorter in the low primaquine arm compared to the high primaquine arm (p=0·0379). When considering all 154 recurrences detected during the 180 days follow-up, including repeated recurrences in a same participant, the median day when any recurrence occurred was 46·0 days (IQR 24·0-74·0, range 10-180), 73·0 days (IQR 47·5-120·0, range 20-180) and 120 days (IQR 73·0-165·0, range 68-180) in the no primaquine, low dose primaquine and high dose primaquine arms, respectively. In the no primaquine arm, the time to any recurrence was not statistically significantly different between randomized (42 days, IQR 18-72, range 12-180) and non-randomized patients (48 days, IQR 29-77, range 10-150, p=0.2956). The time to any recurrence was not statistically significantly different between low dose primaquine arm and high dose primaquine arm (p=0·0742).

All blood-stage infections were treated using artesunate for 7 days at 2 mg/kg/day and arm allocation and primaquine administration were performed after the end of artesunate therapy. Of the 55 patients febrile at enrolment, 93% (51/55) became afebrile within 24h of the first artesunate dose. Parasite clearance was observed for 70% (109/156) of patients within 24h of artesunate initiation, 95% (148/156) were cleared by day 2 and all were parasite-free by day 3 (Appendix Table S6). Parasite clearance of recurrences was similar between the three arms with 87% (87/100), 94% (15/16) and 100% (2/2) of recurrences cleared of parasites within 24h of treatment in the no primaquine, low primaquine and high primaquine arms, respectively. By day 2, 98% (98/100), 100% (16/16) and 100% (2/2) of recurrences were cleared in the no primaquine, low dose primaquine and high dose primaquine arms, respectively. All recurrences were cleared by day 3 in all arms.

No serious AEs (SAE) were reported and all laboratory grade 3 and 4 AEs were asymptomatic and did not require medical attention therefore not meeting the criteria for a SAE. A total of 208 AEs were reported between day 0 and day 90 (table 3). A clinical examination was not performed during the three additional visits at day 120, day 150 and day 180. The proportion of participants encountering an AE in the no primaquine arm (80%, 49/61) was higher than in the low dose primaquine arm (55%, 27/49, p=0·0044) but was not different than in the high dose primaquine arm (67%, 31/46, p=0·1272). There was no statistically significant difference between the two primaquine arms (p=0·2929). The median number of AEs per patient was higher in the no-primaquine arm (2·0, IQR 1·0-2·8, range 0-7) compared to the low dose (1·0, IQR 0·0-1·0, range 0-7, p=0·0001) and the high dose primaquine one (1·0, IQR 0·0-2·0, range 0-4, p=0·0158). There was no difference between the low and high dose primaquine arms(p=0·7231).

Among grade 3 AEs, only one was defined as related to the intervention and was a self-resolved low neutrophil count within 14 days of artesunate treatment upon *P. vivax* recurrence in the no primaquine arm. All other grade 3 AEs were defined unrelated to artesunate or primaquine therapies but most likely resulted from *Plasmodium* infections either at enrolment (fever and anaemia on day 0 and day 1) or upon recurrences (fever and low neutrophil counts prior to artesunate administration). The only patient with a grade 4 AE was in the no primaquine arm at day 7 with a self-resolved low neutrophil count deemed possibly related to the 7-day artesunate regimen.

Primaquine administration was always performed with food intake and was globally well tolerated (Appendix Table S7). Regarding haematological safety, increase in methaemoglobin was more frequent in the high dose primaquine arm (35% of participants, 16/46) compared to the low dose one (10%, 5/49, p=0·0039) but was clinically silent for all participants and resolved after the end of primaquine administration. No patient developed severe anaemia and there was no drop of more than 25% in haemoglobin during primaquine administration.

Mild self-limiting decrease (grade 1) in haemoglobin levels during the 14-day primaquine course were reported in 2 and 7 participants in the low and high primaquine arms respectively. Participants with mild and moderate anaemia were all asymptomatic. The proportion of patients with anaemia during the 90-day follow-up was statistically significantly different for the three arms (p=0·0056) and was higher in the no primaquine arm (48%, 29/61) compared to the low dose arm (20%, 10/49, p=0·0031) and to the high dose arm (26%, 12/46, p=0·0238). There was no statistically significant difference in the proportion of participants with anaemia between the two primaquine regimens (p=0·5120).

## Discussion

This randomised controlled trial provides evidence of higher efficacy to prevent recurrences of the 7·0 mg/kg primaquine regimen compared to the 3·5 mg/kg (total dose, both over 14 days) for infections by *P. vivax* acquired in Cambodia. Our unique study design where (i) patients were relocated during the 90-day follow-up to prevent reinfections, (ii) blood-stage infections were cleared using a short half-life antimalarial (artesunate) and (iii) a frequent and sensitive monitoring of parasite recurrence every second day using both PCR and microscopy, allowed us to evaluate the efficacy of primaquine to prevent *P. vivax* recurrences while minimizing confounding effects for instance through reinfections and the post-prophylactic effects of schizonticidal drugs. By day 90, 7·0 mg/kg of primaquine reduced the risk of *P. vivax* recurrence by 5·5 fold compared to the 3·5 mg/kg regimen. The higher efficacy of high dose primaquine was still apparent at day 180. These results are in line with a recent meta-analysis evaluating efficacy of primaquine dosage.^34^

Using artesunate monotherapy coupled to tight and sensitive follow-up of patients allowed us to document an extremely high rate of relapses occurring in *P. vivax* infected patients when primaquine is not administered, as well as the high number of consecutive relapses, as observed in other studies^27^. These successive relapses inflict a high morbidity to infected individuals as illustrated by the overall higher number of AEs identified in the no primaquine arm compared to the two others with primaquine administration. These AEs are the direct consequence of *P. vivax* relapsing infections and are prevented by primaquine administration. In particular, we show here that successive recurrences in absence of primaquine treatment cause repeated anaemia in infected individuals. Note that this relapse-related morbidity is very likely underestimated in our study because of the frequent follow-up performed: all relapsing infections were re-treated before any clinical consequence of anaemia could be observed, while in real-life settings, repeated anaemia following successive *P. vivax* relapses would have been greater as the disease would have progressed.^5,6,35–39^ Anti-relapse therapy, even with partial efficacy such as the 3·5 mg/kg primaquine regimen, thus provides direct clinical benefit to *P. vivax* infected patients.

Our results confirm that the total dose of 7·0 mg/kg should be the recommended primaquine regimen for individuals with normal G6PD activity in Cambodia and likely in other countries in Southeast Asia. Our results are in line with those of a study published over the course of our trial that compared low dose primaquine (total dose of 3·5 mg/kg as in our study, but over 7 days) efficacy with high dose primaquine (same regimen as we evaluated here) for *P. vivax* infections acquired in Brazil^40^ suggesting that the superior efficacy of 7·0 mg/kg compared to 3·5 mg/kg is likely universal to all tropical *P. vivax* infections. Recently, the WHO issued a recommendation for a shorter course of primaquine over 7 days^14^ to improve compliance, but the total dose of primaquine under this new recommendation is 3·5 mg/kg, the same as the low dose primaquine we evaluated here over 14 days. As efficacy of primaquine is influenced by the total dose administered, rather than by the duration of the treatment or the maximal concentration achieved in the blood (a phenomenon referred to as the total dose effect)^24,41,42^, this new recommendation of 3·5 mg/kg over 7 days is likely insufficient to prevent relapses for infections acquired in Southeast Asia, and probably in most tropical countries^40^. However, 14-day regimens are challenging due to compliance concerns and shorter courses provide clear implementation benefit. A short course (7 days) high dose (7·0 mg/kg total) regimen of primaquine was recently evaluated in a large multicentric clinical trial and was shown to be well tolerated in G6PD normal patients and as effective as 7·0 mg/kg total dose over 14 days^15^.

We have used here 7-day artesunate monotherapy to treat blood-stage infections prior to primaquine administration. While this short half-life antimalarial allowed us to finely characterize any recurrence occurring by not conferring post-treatment prophylaxis, it does not reflect current practices in the vast majority of endemic countries where primaquine is provided concomitantly with treatment with either chloroquine or an artemisinin based combination therapy^14^. These schizonticidal antimalarial drugs will provide longer post-treatment prophylaxis than artesunate due to their longer half-life supressing early relapses.^27,28,43^ However, with perhaps the exception of chloroquine that has been shown to synergize with primaquine in early small studies^44^, but not in larger clinical trials^45^, intrinsic efficacy of primaquine is not affected by the blood-stage drug used and has been shown to be as effective when started after treating blood-stage parasites.^46^

Our study has several limitations. First the sample size is small and the target sample size was not attained for logistical reasons (end of funding period concomitant with decrease in malaria transmission in the study area). Second, this was a single centre open label study conducted in one geographic area of Cambodia. Third, and inherently to *P. vivax* biology, we cannot ascertain that all recurrences observed are relapses and not recrudescence. There is currently no method that can unambiguously tease apart both source of recurrences. We however believe that the majority of recurrences observed are relapses because (i) 95% and 100% of all initial infections were cleared within 48h or 72h of artesunate therapy initiation, respectively, (and confirmed by intensive PCR testing) and (ii) a primaquine dose-dependent decrease in the number of recurrences detected is observed. Further genomic analyses will be performed to provide better insights on these recurrences, providing probabilistic classifications of whether a recurrence is a recrudescence or a relapse.^47^ Finally, our study was performed in tightly controlled conditions, with treatment supervised at all time and sequentially administered with first artesunate and second primaquine and as such does not reflect real life practice and ultimately effectiveness of primaquine.

Overall, our results indicate that a total dose of 7·0 mg/kg of primaquine over 14 days has superior efficacy to prevent *P. vivax* recurrences compared to 3·5 mg/kg of primaquine over 14 days in infections acquired in Cambodia. Both treatments were well tolerated in G6PD normal patients. This is the first study comparing these two regimens in a randomised controlled trial in South East Asia and although more studies should be conducted in other country of the region, our results suggest that countries in South East Asia should endorse the 7·0 mg/kg regimen.

## Data Availability

De-identified patient data will be made available upon reasonable request.

## Contributors

BW, DS and JP conceived the study. VE, SS, LBFD, AO, JS, DS, SH and NK were responsible for data collection. VE, DL, DS and JP oversaw the study. VE, KT, CF, CS, RWvdP, MW and JP did the data analysis. VE and JP wrote the first draft. All authors had full access to all the data in the study and had final responsibility for the decision to submit for publication.

## Data sharing

De-identified patient data will be made available upon reasonable request.

## Acknowledgments

This study was funded by the NIH/NIAID (R01AI146590). J.P is supported by the NIH/NIAID (R01AI173171, R01AI175134 and R61AI187100). MW, BW and JP are supported by the Pasteur International Unit PvESMEE. We are grateful to the SMC members Dr Francois Nosten, Dr Kevin Baird and Dr Jessica Lin for overseeing the safety of this trial.

